# Pattern of Neonatal Outcome of NICU Admissions Born to Teenage Pregnant Women Admitted in a Level-1 Government Hospital from January 2016 to December 2016

**DOI:** 10.1101/2023.02.15.23286000

**Authors:** Arbeen Acosta Laurito

**Affiliations:** Medical Specialist III, General Pediatrics, JR Borja General Hospital

## Abstract

Teenage pregnancy in the Philippines is increasing and alarming. Services in the antenatal care are in accordance with the Department of Health and the World Health Organization serving pregnant women of all ages. Looking at the post-partum women, we will be identifying factors among this specific teenage group who were considered at risk compared to the older pregnant women.

The researcher aims to conduct a study which can give as baseline clinical picture of the product of conceptions among those teenage pregnant women admitted at Justiniano R. Borja General Hospital.

Among the NCU admitted neonates, 120 (20%) were delivered by teenage pregnant women, 131 (21.83%) from 20-24 years old, and 349 (58.17%) from >25 years old pregnant women.

The prevalence of teenage pregnancy in JRBGH was 20%, with 12 years old as the youngest to have live newborn for 2017. Teenage pregnancy ranges from 20 to 23% on monthly delivery census. While NCU babies born to teenage pregnant women ranges from 10 to 26% on monthly census of NCU admission.

This study found out that teenage pregnancy were significantly different from the 20-24 years old in terms of their parity, timing of first ultrasound and hospital expenses. Other maternal profiles which showed no significant difference were; place of residence, menarche, blood pressure, mode of delivery. Hepatitis B status was described by percentage.

Based on this study, parity is significantly different in both age groups. These study population covered the NCU babies. Repeat pregnancies should be considered seriously to prevent more cases of NCU admission or other morbidities. Post-natal care should be accessed and improved in terms of the ante-natal care, tracking system, local networking of referrals, and more improve access to data and data collection for analysis of existing health programs.

The mean hopital expenses is significantly lower in the teenage pregnant women vs 20-24 years old group. The mean hospital expenses for teenage pregnant women is probably higher because of the study population (neonates) were NCU admitted compared to the total delivery. It is noteworthy to mention that majority of these teenage group (study population) are un-employed. And these hospital expenses may correlates to the out-of-pocket expenses from their families or the cost of LGU funding. Thus, the gross cost of hospitalization from these teenage pregnant women would be Php1,044,357.6 (based on the mean hospital expenses x 88 retrieved SOA) or Php 1,424,124 (based on the mean hospital expenses x 120 teenage pregnant women in NCU), or Php 17,635,402 for 2017 (11,867.7 × 1,486 teenage pregnant deliveries).

There were no significant diffrence noted among the neonatal profile variables, namely; birthweight, Apgar score and outcome (complicated or non-complicated). Length of hospital for mothers and length of NCU stay of neonates were found to be not significant in both age groups.

## I. INTRODUCTION

Reducing teenage pregnancy may be viewed in another context. This may be assessed on the impact and risks of teenage pregnancy and associations of risk factors leading to the increase teenage pregnant women population. This study will not aim in reducing teenage pregnancy, but hopefully can enhance more in improving the maternal services for the teenage pregnant women before delivery, and after giving birth.

Policies on antenatal care maybe reviewed and re-focused for these age group so that the family and the health care facilities will be prepared. There is an urgency to assess the needs, to prepare and anticipate extra care for these babies about to be delivered with impending physiologic disabilities.

Birth outcomes may hopefully help convince the public and the teenage population. It will also help us prioritize maternal health services and will help us identify some risk factors that may be related to the increased teenage pregnancy unique to the our population.

The burden of teenage pregnancy will not have a major impact on the mothers’ physiologic and emotional adaptation alone. It will also raised the questions on who will take care of the babies’ needs during the developmental stages and how will these babies survive under a modified family dynamics: unmarried partner, teenage father and extended families.

Various studies were conducted related to teenage pregnancy in different ethnic groups, cultures and in several facility-based study population. It is interesting to note that teenage pregnancy in Cagayan De Oro City and in our facility is increasing.

In 2016, we have 23,043 newborn deliveries (Hospital Database) with an average monthly newborn deliveries of 1,920. We will determine the number and percentage of the NICU admissions among these deliveries born by teenage pregnant women.

The cost classification scheme is dependent on the needs of every program and it is re-classified in this study. This study will focus on the unit financial costs of hospitalization of teenage pregnant mothers and babies’ admissions at NICU. And there will be no program intervention done by the researcher. This study will also assess the costs of admission of these teenage pregnant women which may be too costly compared to their older age group. Categories to be used are the following: medicines (Pharmacy) & supplies (Central Supply Room), laboratory/ ancillary services (Laboratory, Radiology, Ultrasonography) and miscellaneous fees (eg., newborn screening, delivery room services). Professional fees for the medical doctors will be included. But other personnel fees (nurses, nursing attendants, midwives, laboratory technicians) and their services rendered, the buildings (and space) and equipment will not be included in the study.

### General Objective

To determine the neonatal outcome of teenage pregnancy at Justiniano R. Borja General Hospital (JRBGH) from January 2016-December 2016.

### Specific Objectives

1. To determine the profile of teenage pregnant women (19 years old and below) admitted at JRBGH who delivered babies that were admitted at the NICU. Secondary data will be collected from their Admission charts, OPD-Kardex and hospital database as follows:
  a. Age
  b. Residence (urban, rural)
  c. Menarche (years)
  d. Gravidity and parity of pregnancy
  e. Date of first abdominal/vaginal ultrasonography
  f. Hepatitis B status
  g. Systolic BP (admission)
  h. Diastolic BP (admission)
  i. Maternal complications during admission
  j. Length of hospital stay
  k. Unit financial cost of admission
2. To determine the profile of newborns admitted at NICU born to teenage pregnant women, such as:
  a. Gender
  b. Birth weight
  c. Apgar Score (5 minutes)
  d. Type of delivery
  e. Newborn complications
  f. Outcome
  g. Length of hospital stay
  h. Unit financial cost of admission
3. To determine the cost profile (unit financial cost) of hospitalization of the NICU admitted newborns born to teenage pregnant women:
  a. Room and board
  b. Medicine
  c. Supplies
  d. Laboratory
  e. Professional fee
  f. Miscellaneous fees: eg. Delivery room, Newborn screening

### Limitations of the Study

The study population is limited only to the newborns at the Delivery Room who were eventually admitted to the Neonatal Intensive Care Unit (NICU). Thus, babies which were roomed-in immediately, who later became morbid/sick and needs ICU admission, will not be part of the study.

The study is limited to the outcome of neonates among those admitted at the NICU and will not deal on the treatment modalities or management given by the Pediatricians-on-duty.

Data to be collected are secondary data on these retrieved records and thus the circumstances of possible incomplete data will not be known. Moreover, clinical outcome of those babies referred to the other higher level facilities will not be known by the researcher.

Unit financial cost on admission will be based on the attached Statement of Account documents upon discharge. Procedures and services done during prenatal check-ups which were not attached will not be considered.

## II. REVIEW OF RELATED STUDIES

Teenage pregnancy is considered as a community burden noted across the globe and WHO (2014) documented that complications during pregnancy and childbirth were considered as the second cause of death for 15 to 19 year-old girls globally. Every year, some 3 million girls aged 15 to 19 undergo unsafe abortions. Babies born to adolescent mothers face a substantially higher risk of dying than those born to women aged 20 to 24 (WHO, 2014).

Filipino teenage women were described as, “Young, sexually active, and clueless”. The problem here in the Philippines traces its roots to the lack of access to appropriate sexual and reproductive health information and services, the United Nations Population Fund (UNFPA) said (Rodriquez, 2016).

About 16 million girls aged 15 to 19 and some 1 million girls under 15 give birth every year—most in low- and middle-income countries (WHO, 2014). Presently, there are nearly one billion young people aged between 10 and 24 living in the Asia-Pacific region, accounting for more than a quarter of its population. (Rodriquez, 2016). But in the Philippines, it is unique, as reflected in the title of the article by Rodriquez (2016), “Teen pregnancy down in Asia-Pacific, except the Philippines”. The article cited the unpublished analysis of the 2013 Philippine Demographic and Health Survey that “around one-third of adolescent pregnancies were conceived prior to marriage”.

To improve such conditions, the UNFPA suggested the following to the policy makers: support research on sexual and reproductive health; strengthen laws granting the youth access to sexual and reproductive health information, commodities, and services; improve sexuality education; and increase youth participation in policy-making and programming. (Rodriquez, 2016) These steps are acceptable but culturally-variable in terms of its application from country to country.

Access to internet and social media and new information technologies are among the most prominent factors that facilitates early sexual engagement among young Filipinos. As published in the “Regional studies reveal why Filipino millenials engage in early sex”. (Press Release, YAFS 4 Book and Regional Further Studies Launch, 2017).

Some regions in the Philippines showed the significance of education to the sexual and non-sexual behaviors of the young adults. In Cordillera Administrative Region (CAR), young women with low education start childbearing much earlier than the rest of their counterparts. Same finding can be observed in Ilocos Region wherein the proportion of young people who had early sexual initiation is higher among those with low education. Likewise, personal, peer, and community-level factors are more strongly associated than family factors with the youth’s use of ICTs for sex-related purposes in Northern Mindanao. These are just some of the observations among several regions, according to POPCOM Executive Director Dr. Juan Antonio A. Perez III (Press Release, YAFS 4 Book and Regional Further Studies Launch, March 23, 2017).

### Teenage pregnancy prevalence and incidence

Incidence of teenage pregnancy is varied from different countries. The first study to be reviewed is from Saudi Arabia by Abu-Heija et al. (2002) which showed an incidence 0.8%. The study deals on the obstetrics and perinatal outcome of nulliparous teenage pregnant women versus the control group (20-24 years old).

Regional retrospective cohort studies in Thailand (2016) showed teenage pregnancy incidence of 15.24%, comparing to <20 years old women (268) to the 20-34 year old control group (Narukhu, 2016).

Study from India covering 386 teenage pregnant women noted the incident rate at 10% compared with 3,326 older women (Mahavarkar et al, 2008). Still from India, this retrospective study among 620 teenage pregnancies noted an incidence of 4% (Sagili et al, 2012) compared to 14,878 non-teenage women. These two retrospective and comparative studies from India probably differs due to the large population size used by the Sagili et al.

A cross-sectional study by Egbe (2015) in Cameroon from 2009 to 2012 showed prevalence of teenage pregnancy is 13.3%. In a systematic review done by Azevedo (2015) showed an over-all prevalence of 10% in teenage pregnancy. This review included 10 cross-sectional studies and 5 cohort studies. Within this 15 reviewed studies, Brazilian papers showed a higher prevalence at 26%.

### Antenatal visits

Pattern of antenatal visits was documented by Chirayu (2012) by conducting a retrospective study among pregnant women <20 years old which showed low first visit during the first trimester at 17.5% versus 20% in adult pregnant women. Recent study supported the previous claim of low antenatal visits among younger age group, 66.5% vs. adult population at 90.5% (p=0.001) according to the study of Narukhu (2016).

### Mode of delivery NSD

Sagili et al. (2012) showed that normal spontaneous delivery (NSD) is significantly high among 620 teenage pregnant women than 14,878 non-teenage group. A much recent and larger study group by Narukhu (2016), noted that majority of the 957 singleton births were delivered via NSD (59.7%) by the teenage pregnant group compared to adult women (36.4%). However, findings from the study of Pun (2011) showed insignificant relationship between teenage pregnant women delivered NSD (77.4%) comparing to young postpartum women ranges from 20-24 years old (74.6%).

### Maternal Complications

Maternal complications during delivery like cephalopelvic disproportion (CPD) (14.5% vs 26.4%, p=0.001) and postpartum hemorrhage (3.8% vs 8.4%, p=0.016) were lower in teenage pregnant women than the adult group (Narukhu, 2016). According to Chirayu (2012), anemia was considered low at 30.7% from the study group among postpartum teenage women.

Contradicting to the above studies, Pun (2011) showed higher ante-partum hemorrhage among teenage pregnant women (2.4%) compared to the young pregnant group (1.7%). Post-partum hemorrhage rate is much lower at 0.06% versus 0.2% among teenage women and young pregnant, respectively. Both data shows statistically insignificant. Data from Sagili et al. (2012) also showed significant high incidence of anemia and past dates.

Studies among Latin Americans by Azevedo (2015) concluded that adolescent pregnancy is related to increased frequency of maternal and neonatal complications and cesarian section procedures. While Egbe (2015) showed perineal tear at OR-1.66 (95%= level of confidence). There were no reported cases on pre-eclampsia / eclampsia, episiotomy, premature rupture of membrane and ceasarian section. Paper presented by Kirisits (2013) showed seldom complications and no adverse events during pregnancy among 51 Austrian 13-18 years old. The study concluded that social support is necessary for the teenage mom, with adequate interventions both from public and private facilities.

A prospective cross-sectional study (Surval, 2012) among Nepalese teenage pregnant women (<19 yr old) concluded that there were no significant relationships of maternal anemia and hypertension among this study group. While a retrospective study in Kuwait was done to compare <15 years old between >16 years old adolescent pregnancy by Chibber (2014), this study showed a significant increased in the incidence of ectopic pregnancy, pre-eclampsia/eclampsia, PROM, and cesarian section procedures.

### Prematurity

Comparing the two groups of pregnant women, 957 singleton from the study of Narukhu (2016) showed increased risk of prematurity at 16.5% (teenage pregnant) versus the adult group (20-24 years old) at 5.5% (p=0.001). This data were supported from the cohort study of Daniels (2017) which noted an increased rates of preterm and low birth weight among the 10-19 years old, single, nulliparous and uncomplicated NSD. Controlled study by Kovari (2010) among <20 years old versus 20-34 years old pregnant women showed increased risk of prematurity among teenage pregnant group. Same conclusion arrived by Narukhu (2016) using lower age group for the control, 20-24 years old, and significantly supported by Abu-Heija (2002) (p<0.05).

In contrast, previous study in Kathmandu presented that prematurity is not associated with teenage pregnancy (7%) comparing to 20-24 years study group Pun (2011). Prematurity at <37 weeks is at 2.5 (OR, 1.85) at 95% level of confidence from the study of Egbe (2015).

### Low Birth Weight

Covering study group from India, these two retrospective studies showed increased incidence of low birth weight at 13.7% Chirayu (2012) and Sagili et al. (2012). Comparing teenage population versus the control group (20-24 years old), Abu-Heija et al. (2002) documented an increased incidence of low birth weight (p<0.001).

The following reviews does not support that LBW is associated with teenage pregnancy. A cross-sectional study by Pun (2011) compared LBW babies from 15-19 years old (28%) vs 20-24 years old (26.7%) which showed insignificant result. Study by Egbe (2015) showed that LBW is <2.5 (OR, 2.79) at 95% level of confidence.

Moreover, there were no significant relationships of low birth weight and prematurity versus teenage pregnancy among Nepalese women as documented by Surval (2012). But the study showed high perinatal death among married Nepalese at less than <19 years of age.

### Neonatal complications

Apgar Score of <7 is an indicator of neonatal complications (OR=1.66 at 95% level of confidence) among teenage pregnant women based on the study by Egbe (2015). Previous study by Pun (2011) showed insignificant relationship on neonatal complications among teenage pregnant group (17.2%) compared to young pregnant women at 20-24 years old (16.7%). But both studies were contradictory to the study of Kovari (2010) which noted higher neonatal complications versus young pregnant women. Unfortunately, values for this study cannot be retrieved. Chirayu (2012) documented a high neonatal complications at 2.1% if 5-minute Apgar score is <4 compared to 1.2% from babies delivered by young pregnant women (20-24 years old).

### Maternal Health Outcomes

Longitudinal health outcomes presented by Lacobelli (2014a) on adolescent pregnancy. The study significantly showed that rapid repeat pregnancy is higher in teenage group (7.6%) versus the young pregnant women (2.7%). Moreover, case-control study by Lacobelli (2014b) showed that there is less pregnancy-related pathologies but higher psychologic problems from these younger pregnant population. These include suicides, alcohol and drug-abuse, road accidents, and physical abuse.

A more promising strategy to most of the society today is still in its early stage. A randomized controlled study by Taylor (2014) identified a healthier attitudes in reducing teenage pregnancy, as follows: abstain from sex, plans to communicate with parents and increase condom use among teenage group.

### Other determinants

Prospective cross-sectional study in Nepal among the study group of <19 years old showed increased frequency of unbooked cases, low socioeconomic strata, no or inadequate educational background and with little knowledge of contraceptives, less used of contraceptives. Most of the study population were married based on cultural practice (Surval, 2012).

Two conclusive data were drawn from the study of Egbe (2015). The study showed that age and gravidity lend to adverse fetal outcome comparing 14-19 years and 20-29 years old from Cameroon. The other conclusion showed that age/gravidity, unemployment, and marital status would lead to adverse maternal outcome.

### Hospital financial cost

Reviewing the financial cost will provides first look at the effectiveness of *any* health services (Creese and Parker, 1994). Cost were classified into two (2) as Capital Cost and Recurrent Cost from the study of Cresse and Parker (1994). Capital cost includes: vehicles; equipment; buildings/ space; training (non-recurrent); and social mobilization. While recurrent cost includes: personnel involved and its services rendered; supplies; vehicles’ operation and maintenance; training (recurrent); and buildings’ operation and maintenance.

This study will focus on the unit financial cost of hospitalization of the teenage pregnant mother and the baby at the NICU. There is no program interventions done by the researcher. But rather will assess if teenage pregnancy will be more expensive and will add more burden to the family.

By knowing these patterns from other countries, specific problems may be addressed in the community level like Cagayan de Oro City and facility-specific. This will also help us also in the strategic planning for the managers.

For many decades, these consolidated data grossly presents an increasing prevalence of teenage pregnancy worldwide, ranges from 0.8% (Abu-Heija, 2002) in Saudi Arabia to 26% among Brazilian population (from systematic reviews, Azevedo, 2015) as presented in the above studies reviewed.

World Health Organization (2011) recently developed an evidence-based guidelines addressing 6 areas in many low-middle income countries of the world regarding the teenage pregnancy issue, namely: preventing early marriage; preventing early pregnancy through sexuality education; increasing education opportunities and economic and social support programs; increasing the use of contraception; reducing coerced sex; preventing unsafe abortion; and increasing the use of prenatal care childbirth and postpartum care (<u>Chandra-Mouli, 2013).</u>

These six areas need to have a collaborative efforts from several government-private agencies here in our locality, and it requires a mid-term or long-term planning for the government, and few of these areas will be addressed by our health facility as well as JRBGH.

Today, we still do not have baseline data that will consolidate this observation, that the increasing population of teenage pregnancy in Cagayan De Oro City is true. We still have to establish the facts that teenage pregnancy here is really low or high risk to the teenage mothers and the unborn child. The cost of rendering services to these age group may be too costly if we sum it up. Hopefully, by having concrete data, this will be an avenue for more responsive interventions and better actions to achieve our global Sustainable Development Goals (SDG).

## III. METHODOLOGY

### A. Study Design

This is a cross-sectional, analytical and retroactive study to determine the pattern of outcome among neonates admitted at NICU born to teenage pregnant women (13-19 years old).

The study will be conducted in a primary-level government hospital in Cagayan De Oro City from December 2017 to January 2018. The data collection will be performed from December 1 to December 31, 2016 and the second month will be used for the data analysis.

The study methodology will be submitted and subject for approval. The primary investigator is permanently employed as a Pediatrician in JRBGH.

A final copy of the study will be given to the Chief of Hospital, department chairmen from the Pediatric and OB-Gyne Department. The participants included in the study will not be informed and will not be given a copy of the study’s outcome.

The written permissions to conduct the study will be attached as an APPENDIX and will be secured from the Chief of Hospital, Chief of Clinics, and NICU Nurse Manager. Permissions to collect the data from the Medical Record In-Charge (Patients Charts) and the OPD Nurse Manager (Kardex/OPD Chart) will also be attached. Data collected from these charts will be verified from the hospital database by the IT in-charge.

### B. Study Population

This is a retroactive and cross-sectional study based on NICU admission logbook and patients’ medical records. Thus, the sample size will be determined by the number of teenage pregnant women (13-19 years old) who delivered babies and eventually admitted at the NICU.

The research will utilize retrieval of secondary data from the NICU logbook, Medical Records, OPD-Kardex and hospital database. There will be no interviews done to the participants written in the logbook nor the personnel in-charge. The data to be collected are personal data without the participant’s specific consent required. Since there will be no direct participants’ involvement during the study, informed assent/ consent will not be needed.

Patients’ data will be handled with strict confidentiality. NICU admission logbook will be checked from the hospital database (accessed by IT in-charge only) for retrieval of babies’ and mothers’ complete records.

Data from these babies who were born to teenage pregnant women will be collected, and respective Admission and OPD charts will be retrieved. Then the retrieved data will be encoded in spreadsheet format and anonymized by the researcher before subjected to statistic analysis. Names of the mothers will be replaced by codes (random numbers). Names of the corresponding babies will be labeled based on numbers (Bb#1, Bb#2).

Codes will be decoded only by the researcher after the analysis of the study to counter-check the entries which may be altered along the study course. Functional anti-virus computer software will also be updated and files backed up for security reasons.

There will be no noted potential risk to the participants. While the potential risk for the researcher is on the the chart retrieval with possible exposure to dust (from old charts), eye straining during data validation, and/ or when data will be accidentally deleted. Personnel involved during the charts/data retrieval will be advised of these possible risks. These risks are minimal and can be managed by protecting oneself and anticipation along the course of the study.

### C. Study Tools

1. To determine the profile of teenage pregnant women (19 years old and below) admitted at JRBGH who delivered babies that were admitted at the NICU. Secondary data will be collected from their Admission charts, OPD-Kardex, and hospital database as follows:
  a. Age
  b. Residence (urban, rural)
  c. Menarche
  d. Gravidity and parity of pregnancy
  e. Date of first abdominal ultrasonography
  f. Hepatitis B status
  g. Systolic BP on admission
  h. Diastolic BP on admission
  i. Maternal complications
  j. Length of stay (LOS)
  k. Unit financial cost of admission
2. 2. To determine the profile of newborns admitted at NICU born to teenage pregnant women, such as:
  a. Gender
  b. Birth weight
  c. Apgar Score (5 minutes)
  d. Type of delivery
  e. Newborn complications
  f. Outcome
  g. Length of stay (LOS)
  h. Unit financial cost of admission

### D. Inclusion Criteria

1. All NICU admissions irregardless of the outcome born from January 2016 to December 2016.
2. All NICU admissions with retrieved Admission charts.
3. All NICU admissions born to teenage pregnant women (19 years old and below) irregardless of their ward admission.
4. All teenage pregnant women with babies admitted at NICU (≤19 years old) with Admission charts, OPD-Kardex, and verified records from hospital database.
5. All NICU admissions born to teenage pregnant woman irregardless of the mode of delivery: caesarian section or NSD.

### E. Exclusion Criteria

1. Teenage pregnant woman admitted at JRBGH with gynecologic problem/s, who delivered an abortion and/ or stillbirth is excluded from the study.
2. Teenage pregnant woman with either no admission chart or OPD-Kardex retrieved.
3. NICU admissions with no admission charts retrieved.
4. Newborns from the Delivery Room immediately room-in to the ward.
5. Transit/Home delivery babies seen at the Delivery room, OB-Emergency room and Emergency room are excluded.

## IV. RESULTS

A total of 251 newborn babies admitted at the NCU born to teenage pregnant women age (120, 12-19 years old) and 20-24 years old pregnant women (131) were used in the study. It was conducted at LGU-owned level 1 public hospital in Cagayan De Oro CIty from January to December 2017.

There were 2,932 deliveries from 2013 and doubled in 2016 at 6,715. Figure 4.1 demostrated the increasing yearly census of deliveries for the span of 5 years from 2013 to 2017. A detailed summary of monthly census in Table 4.1, showed that there were 6,942 total deliveries of pregnant women at JRBGH as of January 2017 to December 2017.

**Table 4.1.** Monthly census of all pregnant women who were admitted at JRBGH from January to December 2017.

| Age | JAN | FEB | MAR | APR | MAY | JUN | JUL | AUG | SEP | OCT | NOV | DEC | TOTAL | % |
| --- | --- | --- | --- | --- | --- | --- | --- | --- | --- | --- | --- | --- | --- | --- |
| 12-19 | 125 | 91 | 125 | 124 | 128 | 114 | 127 | 131 | 138 | 119 | 135 | 129 | 1,486 | 21.40 |
| 20-24 | 168 | 144 | 161 | 192 | 180 | 181 | 159 | 175 | 195 | 147 | 177 | 187 | 2066 | 29.77 |
| >25 | 269 | 206 | 251 | 295 | 307 | 278 | 290 | 294 | 292 | 279 | 293 | 336 | 3,390 | 48.83 |
| Total | 562 | 441 | 537 | 611 | 615 | 573 | 576 | 600 | 625 | 545 | 605 | 652 | 6,942 | 100 |

**Figure 4.1.**
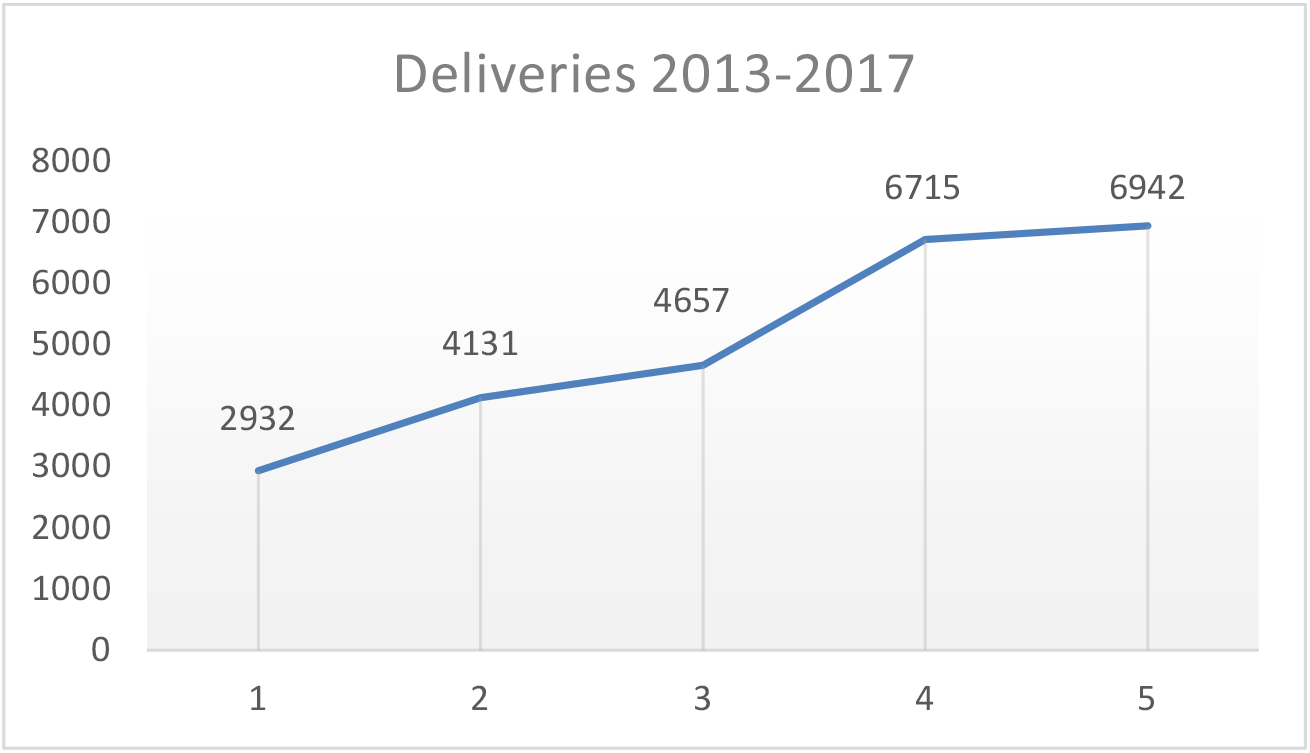
Yearly Census of Deliveries Among Pregnant Women Admitted at JRBGH from 2013 to 2017.

This table also presented that majority of mothers who gave birth (48.83%) were above 25 years of age, followed by the 20-24 years age group (29.77%) and the teenage mothers (21.4%).

The admission of teenage pregnant women (12-19 years old) peaked on September and November, and lowest on February (91) of 2017. For the adult group (20-24 years old), the admission peaked on the month of September (195) and April (192), and the lowest admission was on the month of February with 144 deliveries. The youngest age who gave birth was 12 years old (see Table 4.2) and the oldest was 47 years old.

**Table 4.2.** Monthly census of all teenage pregnant women (≤19 years old) who were admitted at JRBGH from January to December 2017.

| Age (years) | JAN | FEB | MAR | APR | MAY | JUN | JUL | AUG | SEP | OCT | NOV | DEC | TOTAL | % |
| --- | --- | --- | --- | --- | --- | --- | --- | --- | --- | --- | --- | --- | --- | --- |
| 12 | 1 | 0 | 0 | 0 | 0 | 0 | 0 | 0 | 0 | 0 | 0 | 0 | 1 | 0.01 |
| 13 | 1 | 1 | 0 | 0 | 0 | 0 | 0 | 1 | 0 | 0 | 0 | 0 | 3 | 0.04 |
| 14 | 3 | 1 | 1 | 2 | 4 | 0 | 1 | 2 | 1 | 2 | 0 | 4 | 21 | 0.30 |
| 15 | 4 | 2 | 12 | 6 | 5 | 7 | 10 | 7 | 9 | 8 | 11 | 5 | 86 | 1.24 |
| 16 | 13 | 17 | 15 | 14 | 8 | 6 | 24 | 13 | 15 | 16 | 22 | 18 | 181 | 2.61 |
| 17 | 16 | 18 | 22 | 27 | 29 | 32 | 30 | 28 | 40 | 32 | 30 | 34 | 338 | 4.87 |
| 18 | 50 | 26 | 43 | 34 | 41 | 29 | 32 | 49 | 36 | 32 | 42 | 33 | 447 | 6.44 |
| 19 | 37 | 26 | 32 | 41 | 41 | 40 | 30 | 31 | 37 | 29 | 30 | 35 | 409 | 5.89 |
| **Total** | 125 | 91 | 125 | 124 | 128 | 114 | 127 | 131 | 138 | 119 | 135 | 129 | 1,486 | 21.4 |

Table 4.2 shows the distribution of age of teenage pregnant women (≤19 yo) admitted for the year 2017. The youngest was 12 years old who delivered on the month of January. Teenage pregnant women census was 1,486 which comprises the 21% of the total admitted population of pregnant women. Majority of the teenage pregnant women were at the age of 18 years old (6.44%), followed by 19 years old (5.89%), 17 years old (4.87%). Starting from the age of 17 years old, the census of deliveries decreases until the age of 12 years old.

Table 4.3 shows the age distribution of 20-24 years old pregnant women who were admitted at JRBGH from January to December 2017. This group comprises 30% (2,066) of all pregnat women who delivered babies in 2017. Majority of these of pregnant women belongs to age 21 years old (6.34%) but the difference is not widely varied as compared to the rest of the age groups which ranges from 5.29% to 6.14%.

**Table 4.3.**
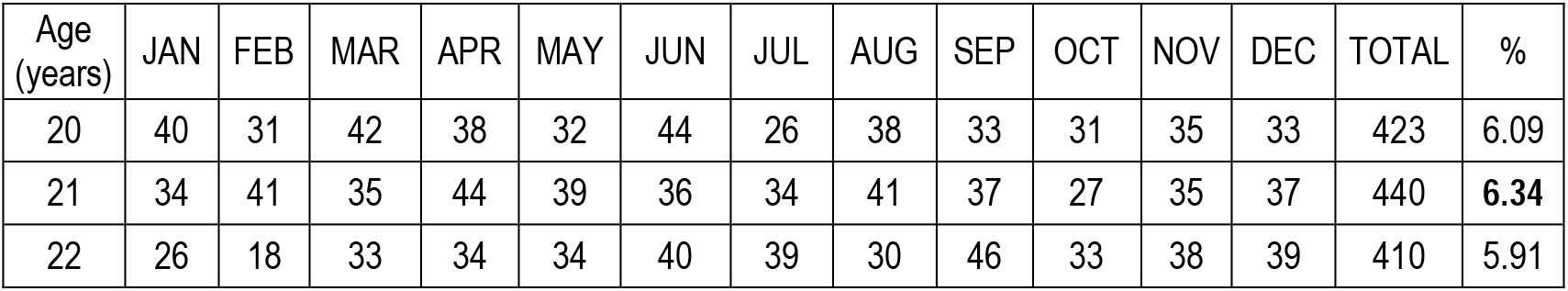
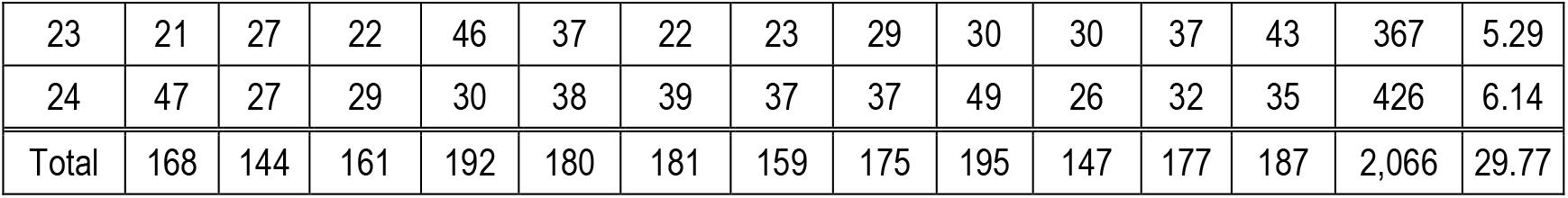
Monthly census of pregnant women age 20-24 years old who were admitted at JRBGH from January to December 2017.

There were 600 NCU admissions from January to December 2017 based on NCU logbooks. One hundred twenty (20%) newborns were born to teenage pregnant women while 131 (21.83%) were born to 20-24 years old pregnant women. These were the study population used for the analysis of the study. Majority of these NCU admissions were born to mothers ≥ 25 years old (58.17%). (see Table 4.4). NCU admission peaked at months of April and May with 62 and 63 admissions, respectively.

**Table 4.4.**
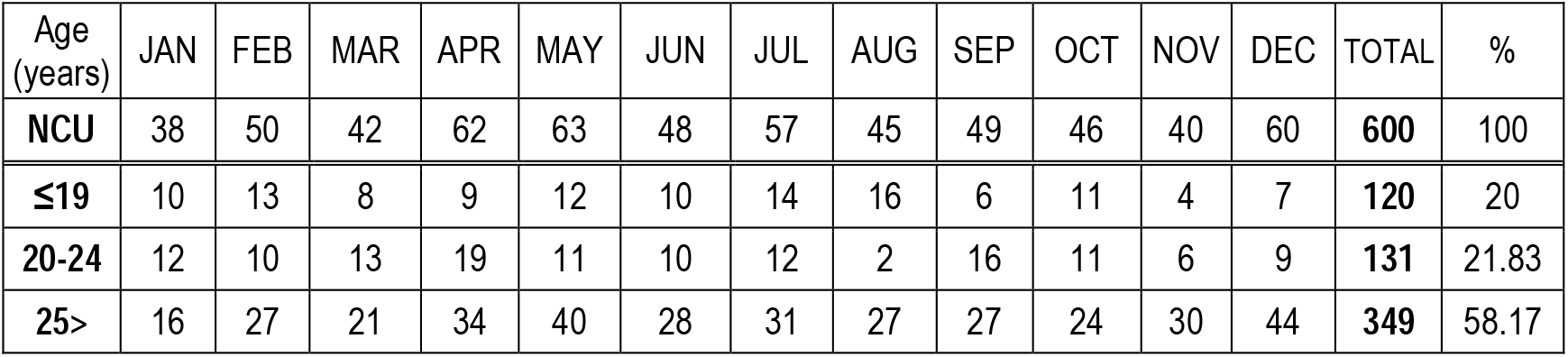
Monthly census of newborns admitted at NCU born to pregnant women (all ages) who were admitted at JRBGH from January to December 2017.

Above Table 4.5, showed the detailed monthly distribution of age ranges from 12 to 19 years old teenage pregnant women (120) with babies admitted at NCU. This is a sub-group among the 600 NCU admission. Majority of these teenage pregnant women is 18 year-old with 37 NCU babies, followed by the 17 year old group. One NCU babies each for 12 and 13 year-old women. The month of August had the most number of NCU admission (16) from these teenage pregnant women.

**Table 4.5.**
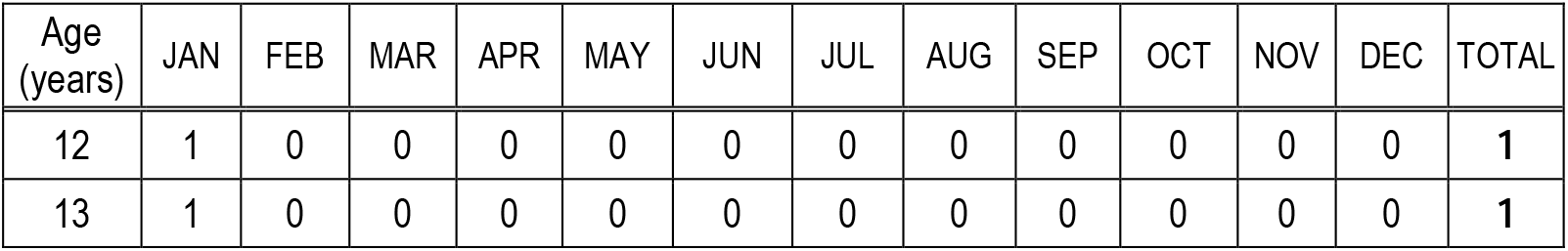
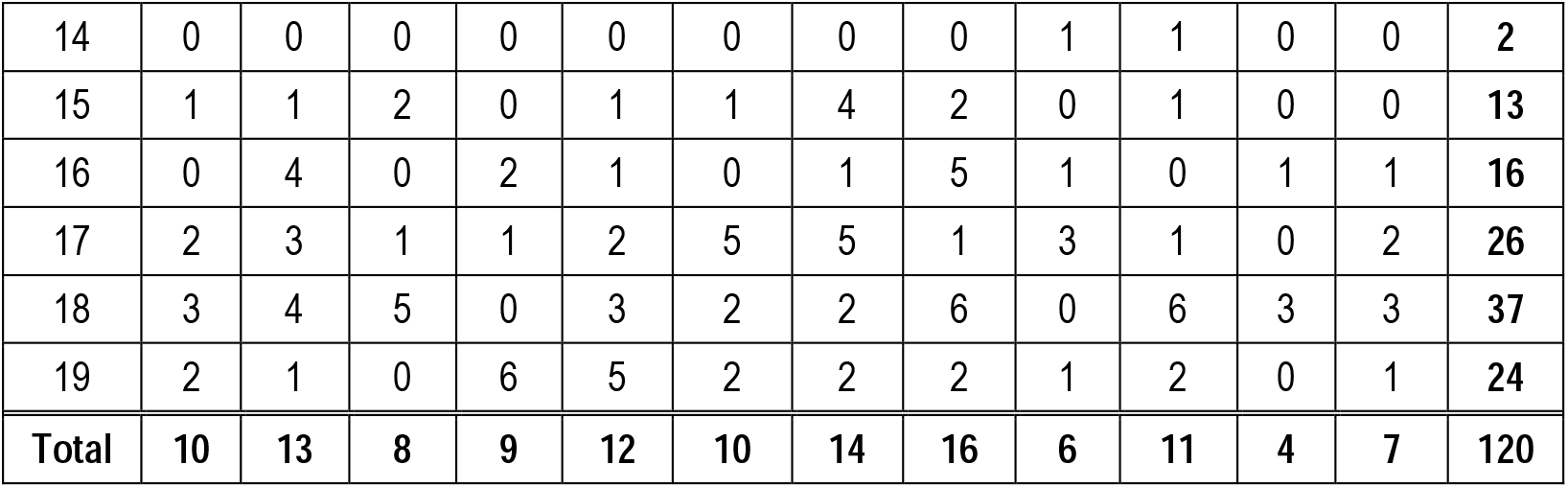
Age distribution of <u>teenage pregnant women</u> who delivered babies admitted at NCU from January to December 2017 (n=120).

The monthly distribution of age ranges from 20 to 24 years adult pregnant women with 131 babies admitted at NCU. This is a sub-group among the 600 NCU admission (21.83%). Among this group, the 21 year-old women had the most number of NCU admission (31), followed by the 22 year-old group (29). In contrast to the teenage group, the month of April had the most number of NCU admission (19) from these adult pregnant women (Table 4.6).

**Table 4.6.** Age distribution of <u>20-24 years old pregnant women</u> who delivered babies admitted at NCU from January to December 2017 (n=131).

| Age (years) | JAN | FEB | MAR | APR | MAY | JUN | JUL | AUG | SEP | OCT | NOV | DEC | TOTAL |
| --- | --- | --- | --- | --- | --- | --- | --- | --- | --- | --- | --- | --- | --- |
| 20 | 4 | 1 | 1 | 4 | 4 | 2 | 0 | 0 | 3 | 0 | 1 | 1 | 21 |
| 21 | 3 | 2 | 6 | 4 | 3 | 2 | 3 | 1 | 0 | 2 | 3 | 2 | 31 |
| 22 | 0 | 1 | 3 | 5 | 2 | 3 | 4 | 0 | 6 | 3 | 1 | 1 | 29 |
| 23 | 1 | 4 | 2 | 3 | 1 | 1 | 3 | 1 | 3 | 5 | 0 | 3 | 27 |
| 24 | 4 | 2 | 1 | 3 | 1 | 2 | 2 | 0 | 4 | 1 | 1 | 2 | 23 |
| Total | 12 | 10 | 13 | 19 | 11 | 10 | 12 | 2 | 16 | 11 | 6 | 9 | 131 |

Objective No. 1: To determine the profile of teenage (19 years old and below) and the control (20-24 years old) pregnant women admitted at JRBGH who delivered babies that were admitted at NCU. Secondary data will be collected from their Admission charts and hospital database as follows:

l Age
m Residence (urban, rural)
n Menarche (years)
o Parity of pregnancy
p Timing of first ultrasonography (abdominal/vaginal)
q Blood Pressure on admission: hypertensive or non-hypertensive
r Length of stay (LOS)
s Mode of delivery: NSD, cesarian section
t Hepatitis B status
u Maternal complications during admission

Among the pregnant women in the study population, majority were residing in the urban barangays both from teenage pregnant women and adult group with 77 and 86, respectively. Consistently, the most number of pregnant women came from where the facility is located, Barangay Carmen, for both age groups.

For the city residents, Barangay Lumbia had 11 admitted women, followed by Brgy. Cugman (7), and 6 admissions each from Brgys. Balulang, Camaman-an, Kauswagan, and Canito-an. From neighboring municipalities, Talakag (Bukidnon) had 16 admitted women, then Opol (13, Misamis Oriental), and Baungon (5, Bukidnon) (see Table 4.7).

**Table 4.7.** Distribution of residence as urban and rural among teenage and 20-24 years old pregnant women (January to December 2017).

| Residence |  | < 19 yo (n=119) |  | 20-24 yo (n=130) |
| --- | --- | --- | --- | --- |
| Classification | Urban | 77 | Urban | 86 |
|  | Rural | 42 | Rural | 44 |
| Distribution<br>(most<br>common) | Carmen | 13 | Carmen | 12 |
|  | Talakag | 8 | Talakag | 8 |
|  | Opol | 7 | Cugman | 7 |
|  | Kauswagan | 6 | Balulang | 6 |
|  | Lumbia | 5 | Camaman-an | 6 |
|  | Baungon | 5 | Canito-an | 6 |
|  |  |  | Lumbia | 6 |
|  |  |  | Opol | 6 |

Table 4.8 showed that majority of the pregnant women had their menarche less than 13 years of age both for teenage and 20-24 years old group with 69 and 66, respectively. Followed by the population having their menarche beyond 14 yo, 27 for teenage group, and 36 for the 20-24 years old group.

**Table 4.8.**
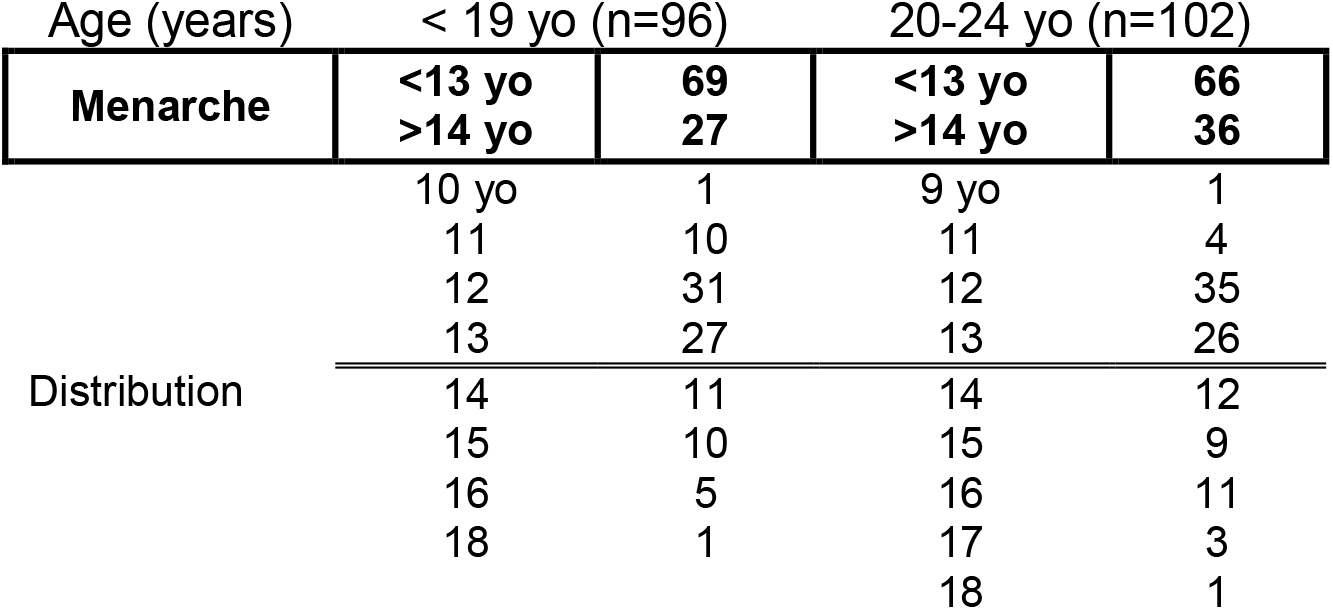
Distribution of teenage and control group pregnant women according to the their menarche (years) (January to December 2017).

The earliest menarche for the teenage pregnant women’s group was 10 years old while younger at 9 years old for the control group. Majority of the study population for both age group had their menarche at 12 years of age.

Majority of the teenage pregnant women were nulliparous during their admission (87) and decreases in frequency with increasing numbers of parity. Meanwhile, 65 pregnant women from the 20-24 years old age group were nulliparous during admission, with decreasing frequency as parity increases comparable to the teenage group. The highest parity of pregnancy for teenage women is 4 and 5 or the older group (Table 4.9).

**Table 4.9.**
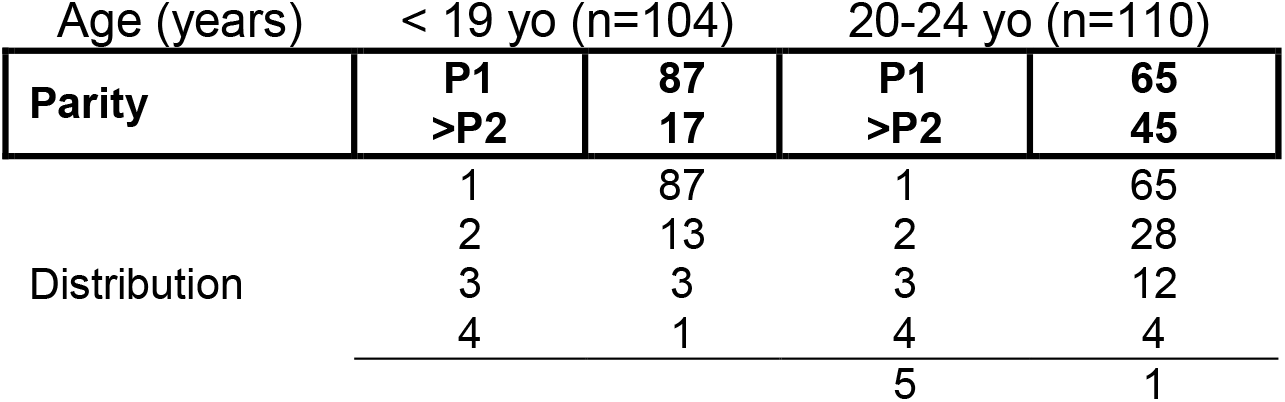
Distribution of teenage and 20-24 years old pregnant women according to their parity of pregnancy (January to December 2017).

| Age (years) |  | < 19 yo (n=104) |  | 20-24 yo (n=110) |  |
| --- | --- | --- | --- | --- | --- |
| Parity |  | P1 | 87 | P1 | 65 |
|  |  | >P2 | 17 | >P2 | 45 |
| Distribution | 1 |  | 87 | 1 | 65 |
|  | 2 |  | 13 | 2 | 28 |
|  | 3 |  | 3 | 3 | 12 |
|  | 4 |  | 1 | 4 | 4 |
|  |  |  |  | 5 | 1 |

Ultrasonography (UTS) procedures were requested and performed either from JRBGH, or outside of the facility from the private clinic or barangay health facilities. Some had series of ultrasound procedures but this study deals only with the timing of the first ultrasonography. The study population were group, as follows: 1st trimester for the 1st week of pregnancy to 12th weeks, 13 to 27 weeks for 2nd trimester, and 28 to 42 weeks for the 3rd trimester. Approximately, 40% from each group do not have ultrasound results attached to their medical records or requested but not done at all.

Majority of teenage pregnant women (56) had their ultrasound performed on the 3rd trimester of pregnancy (mode= 29 weeks) compared to those who had their UTS at the 2nd trimester (16) and none for the 1st trimester (see Table 4.10).

**Table 4.10.** The timing of ultrasound (abdominal/vaginal) among teenage and 20-24 years old pregnant women admitted at JRBGH from January to December 2017.

| Trimester (weeks) | < 19 yo<br>(n=72/120) 60.0% | 20-24 yo<br>(n=81/131) 61.8% |
| --- | --- | --- |
| **1st trimester (1-12 wks)** | **0** | **4** |
| **2nd trimester (13-27 wks)** | **16** | **31** |
| **3rd trimester (28-42 wks)** | **56** | **46** |
| Mean weeks | 25.22 | 22.03 |
| Mode | 29 | 27 |
| Early | 13 | 8 |
| Late | 38 | 38 |

Likewise, the 20-24 year old pregnant women had congruent pattern with the first group. Majority had their UTS at the 3rd trimester of pregnancy (46), then followed by 31 at 2nd trimester and 4 had their UTS during their 1st trimester of pregnancy. The mode (27 weeks) is earlier than the teenage group.

Moreover, the mean weeks of UTS in teenage women (25 weeks) is much later than the 20-24 years old group at 22 weeks. The earliest possible UTS is also late in the teenage women, 13 weeks of pregnancy, compared to 8 weeks from the 20-24 years old group.

Based on charts review, data from Table 4.11 showed that majority of the study population were non-hypertensive during pregnancy and after delivery for both systolic and diatolic blood pressures. Among the teenage women (97), 11 were hypertensive and 86 were non-hypertensive, while 16 were hypertensive among 20-24 years old women and 86 noted not to have elevated blood pressure.

**Table 4.11.** Classification of blood pressure (systole and or diastole) of teenage and 20-24 years old pregnant women admitted at JRBGH from January to December 2017.

| Gestational HPN | < 19 yo (n=97) | 20-24 yo (n=102) |
| --- | --- | --- |
| Hypertensive | 11 | 16 |
| Non-hypertensive | 86 | 86 |

Table 4.12 presented the length of hospital stay (LOS) among teenage and 20-24 years old postpartum women. Among 98 teenage women, the mean LOS is 5.16 days while 5.54 days among 20-24 years old women. The median days for teenage and 20-24 years old are 5.5 days and 6 days, respectively.

**Table 4.12.** Length of hospital stay (LOS) of teenage and 20-24 years old postpartum women admitted at JRBGH from January to December 2017.

| LOS (days) |  | < 19 yo (n=98) |  | 20-24 yo (n=107) |
| --- | --- | --- | --- | --- |
| Mean LOS |  | 5.16 |  | 6.54 |
| Median |  | 5.5 |  | 6 |
| Mode |  | 5 |  | 7 |
| Distribution | <24 hrs | 3 | <24 hrs | 5 |
|  | 1 | 2 | 1 | 1 |
|  | 2 | 7 | 2 | 4 |
|  | 3 | 7 | 3 | 5 |
|  | 4 | 13 | 4 | 14 |
|  | **5** | **16** | 5 | 11 |
|  | 6 | 17 | **6** | **21** |
| Longest day | 20 days | 1 | 22 days | 1 |

The mode of LOS is 5 days for teenage group (16 women) while 6 days for the 20-24 years old group (21 women). The longest LOS (days) is 22 days for the 20-24 years old group which is longer compared to 20 days only from the teenage group.

Majority of the 119 teenage pregnant women had normal spontaneous deliveries-NSD (95) while 24 had their delivery through cesarian section (1 for cesarian section due to twin pregnancy). There was one (1) each for both forcep delivery and twin-NSD delivery.

Table 4.13 also showed that majority of the 20-24 years old pregnant women (129) had NSD (93) and only 32 delivered through cesarian section. There were 4 assisted deliveries (forcep delivery), none for twin pregnancy for this age group.

**Table 4.13.**
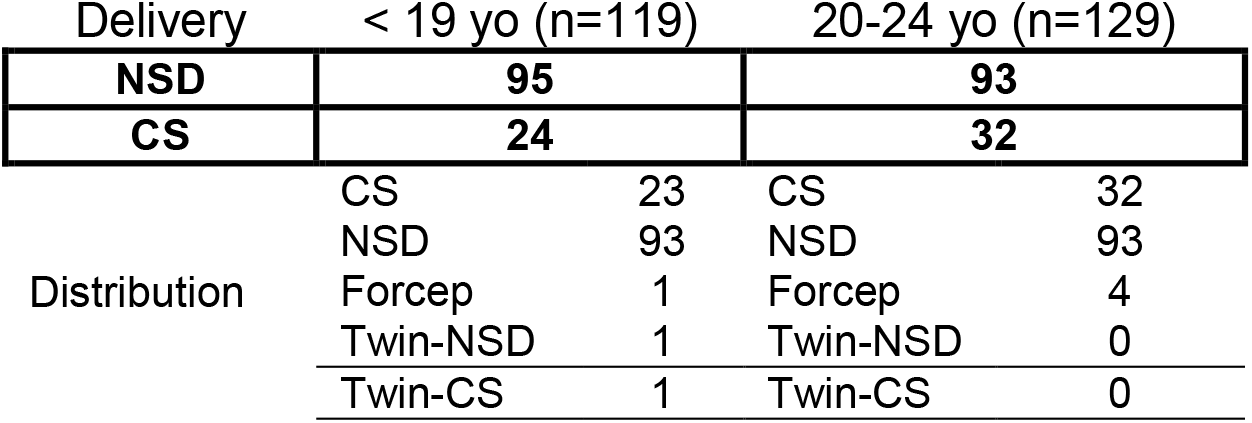
Mode of delivery for teenage and 20-24 years old pregnant women admitted at JRBGH from January to December 2017.

Based on charts review, among the 91 teenage pregnant women with laboratory results attached, one (1.1%) tested Hepatitis B reactive mother and majority were non-Hepatitis B reactive (90, 98.9%). Moreover, the 129 20-24 years old group, same trend was noted, majority were non-Hepatitis B reactive (97, 96%) and there were 4 (4%) tested Hepatitis-B reactive (Table 4.14).

**Table 4.14.**
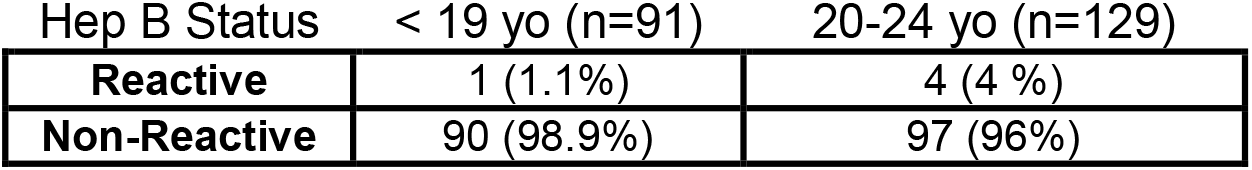
Hepatitis B status of teenage and 20-24 years old pregnant women admitted at JRBGH from January to December 2017.

| Hep B Status | < 19 yo (n=91) | 20-24 yo (n=129) |
| --- | --- | --- |
| **Reactive** | 1 (1.1%) | 4 (4 %) |
| **Non-Reactive** | 90 (98.9%) | 97 (96%) |

Table 4.15 showed 9 teenage pregnant women (9/119=7.6%) had maternal complications during delivery. Three (3) had anemia and had blood transfusions with 1 unit of blood component each. There were two (2) women with multifetal pregnancy (2/119). One case each for pneumonia, malposition, preeclampsia and thrombocytopenia.

**Table 4.15.** Maternal complications of teenage and 20-24 years old pregnant women during admission at JRBGH from January to December 2017.

| < 19 yo (n=119) |  |  | 20-24 yo (n=129) |  |  |
| --- | --- | --- | --- | --- | --- |
| Case | Frequency | % | Case | Frequency | % |
| Anemia | 3 | 33.3 | Anemia | 4 | 26.6 |
| Multifetal | 2 | 22.2 | Anemia +<br>eclampsia | 1 | 6.7 |
| Pneumonia | 1 | 11.1 | CPD | 2 | 13.3 |
| Malposition | 1 | 11.1 | PROM | 3 | 20 |
|  |  |  | Preeclampsia | 2 | 13.3 |
| Preeclampsia | 1 | 11.1 | Pneumonia,<br>pleural effusion | 1 | 6.7 |
|  |  |  | Severe<br>preeclampsia | 1 | 6.7 |
| Thrombocytopenia | 1 | 11.1 | UTI | 1 | 6.7 |
| Total | 9 | 99.9 | Total | 15 | 100 |

Several studies summarizes maternal complications among teenage pregnant women. Khier et al, 2017 study revealed that 7 (7%) of the control group and 18 (18%) of the case group had medical problems during pregnancy with significant differences between the two groups (p value = 0.011). Risks of placenta previa, postpartum haemorrhage > 1000 ml and perineal rupture were significantly lower among teenagers (Tyrberg et al., 2013). But intrapartum (CPD, fetal distress) and postpartum complications (PPH, retained placenta, puerperal infection) were similar between teenage and adult mothers (Kovavisarach et al, 2010). Furthermore, adolescent mothers had a higher incidence of pre-eclampsia (odds ratio [OR] 2.017 95% confidence interval [CI]: 1.045–3.894, *P*=0.03), preterm deliveries (OR: 1.655, 95% CI: 1.039–2.636, *P*=0.03) (Medhi et al, 2016).

Objective No. 2 To determine the profile of newborns admitted at NCU born to teenage pregnant women and the control group, such as:

a. Sex
b. Birth weight (kg)
c. Apgar Score (first 5 minutes)
d. Outcome: complicated or non-complicated
e. Newborn complications
f. Length of NCU stay (LOS), days

Table 4.16 shows the distribution of NB according to their sex. Majority of these babies are male (74, 61.7%) and there were 46 (38.3%) females among the 120 babies delivered by teenage women. While, there were 83 males (63.4%) and 48 females (36.6%) among 131 NB delivered by 20-24 years old group from January to December 2017.

**Table 4.16.** The sex distribution of NCU admitted babies born to teenage and 20-24 years old pregnant women.

| Sex | < 19 yo (n=120) |  | 20-24 yo (n=131) |  |
| --- | --- | --- | --- | --- |
| Male | 74 | 61.7% | 83 | 63.4% |
| Female | 46 | 38.3 | 48 | 36.6 |
| Total | 120 | 100 | 131 | 100 |

The mean birth weight for 121 babies born to teenage women is 2.507 kgs (median = 2.6 kgs) while mean birth weight among 131 babies born to 20-24 years old women is 2.649 kgs (median = 2.75 kgs). In both groups, the mode of the birth weight is 3 kgs.

The recommended birth weight is 2.5 kgs for newborn. Table 4.17 showed that majority (63) of the 121 NB born to teenage women weighed ≥2.5 kgs and 52 babies were ≤ 2.4 kgs. Five (5) NB weighed more than 3.5 kgs with 4.45 kgs as the heaviest weight while the smallest weight was 0.65 kgs.

**Table 4.17.** The birth weight of NCU admitted babies born to teenage and 20-24 years old pregnant women.

| Birth weight (kgs) |  | < 19 yo (n=121) |  | 20-24 yo (n=131) |
| --- | --- | --- | --- | --- |
| **≤ 2.4 kgs** |  | **52** |  | **41** |
| **≥ 2.5 kgs** |  | **63** |  | **76** |
| Distribution | <1.0 | 4 | <1.0 | 0 |
|  | 1.0-1.95 | 27 | 1.0-1.95 | 23 |
|  | 2.0-2.4 | 22 | 2.0-2.4 | 18 |
|  | 2.5-3.4 | 66 | 2.5-3.4 | 76 |
|  | >3.5 | 5 | >3.5 | 14 |
| Mean |  | 2.507 |  | 2.649 |
| Median |  | 2.6 |  | 2.75 |
| Mode |  | 3 |  | 3 |
| Heaviest |  | 4.45 |  | 4.45 |
| Lowest |  | 0.65 |  | 0.6 |

There were 76 NB weighed ≥2.5 kgs (among 131 NB) born to 20-24 years old women and only 41 babies were ≤ 2.4 kgs. The heaviest NB weighed at 4.45 kg and the smallest was at 0.6 kg. Fourteen (14) NB weighed ≥3.5 kgs as large-for-gestational (LGA) babies.

Apgar score for the newborn were taken at first 5-minutes of life and the next 10-minutes of life. The first 5-minute is critical since it will help in the decision to initiate appropriate resuscitative management which is use in this study.

Majority of the 121 babies born to teenage women, had Apgar score of ≥ 7 (79 babies) while 42 babies with score ≤ 6. The same pattern is true to 131 babies born to 20-24 years old women with majority (82) had an Apgar score of ≥ 7, while 49 babies had an Apgar score of ≤ 6. The mean Apgar score for both teenage and 20-24 years old were 6.748 and 6.636, respectively. (Table 4.18).

**Table 4.18.** Apgar Score (first 5 minutes) of NCU admitted babies born to teenage and 20-24 years old pregnant women.

| Apgar Score | < 19 yo (n=121) |  | 20-24 yo (n=131) |  |
| --- | --- | --- | --- | --- |
| **≤ 6** | **42** |  | **49** |  |
| **≥ 7** | **79** |  | **82** |  |
| Distribution | 0-3 | 25 | 0-3 | 24 |
|  | 4-6 | 17 | 4-6 | 25 |
|  | 7-8 | 8 | 7-8 | 18 |
|  | 9-10 | 71 | 9-10 | 64 |
| Mean | 6.748 |  | 6.636 |  |
| Median | 9 |  | 8 |  |
| Mode | 9 |  | 9 |  |

Among the 109 babies delivered by teenage women, 22 were categorized as complicated deliveries and 87 were non-complicated. The 22 complicated cases comprise the expired babies (17), IUFD (1), and 4 transferred babies to other facilities for tertiary-level management. There were 78 babies roomed-in with their mothers, and discharged home (9) from NCU which composed the 87 non-complicated deliveries (Table 4.19).

**Table 4.19.**
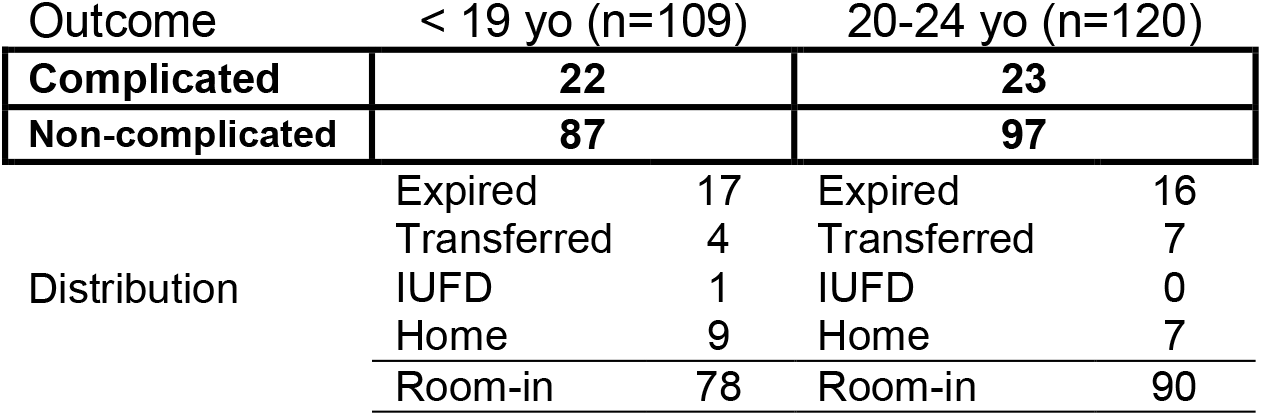
The clinical outcome of NCU admitted babies born to teenage and 20-24 years old pregnant women.

Table 4.19 shows 120 babies delivered by 20-24 years old women, and most (97) of the babies were delivered with no complications. This consist the following: roomed-in (90) with their mothers and discharged home (7). The 23 complicated cases, were as follows: 16 expired due to medical conditions and 7 babies were transferred for further management at the tertiary-level facility. No case for IUFD.

Among babies born to teenage pregnant women, the mean LOS is 3.314 days. Majority (31 babies) stayed in the NCU for 24 hours, followed by the following: 2 days – 27 babies, 3 days - 13 babies, and 12 babies stayed less than 24 hours at NCU. The longest stay in NCU was at 40 days (Table 4.20).

**Table 4.20.** The length of stay (LOS) of NCU admitted babies born to teenage and 20-24 years old pregnant women.

| LOS (days) | < 19 yo (n=105) |  | 20-24 yo (n=120) |  |
| --- | --- | --- | --- | --- |
| Mean LOS | 3.314 |  | 3.175 |  |
| Median | 2 |  | 2 |  |
| Mode | 1 |  | 2 |  |
| Distribution | <24 hrs | 12 | <24 hrs | 20 |
|  | 1 | 31 | 1 | 20 |
|  | 2 | 27 | 2 | 26 |
|  | 3 | 13 | 3 | 16 |
|  | 18 days | 1 | 19 | 1 |
|  | 33 days | 1 | 20 | 1 |
|  | 40 days | 1 | 22 | 1 |

The mean LOS is 3.23 days for babies delivered by 20-24 years old women and majority (26 babies) stayed at NCU for 2 days. These were followed by 20 babies for both less than 24 hours (0 day) and 1 day. The longest stay in NCU was at 22 days. (Table 4.20).

There are 48 NCU admission born to teenage pregnant women with complications. Majority had respiratory complications (15) followed by infectious in etiology, neurologic (6) an others (6) (Table 4.21 below). Meanwhile, there were 65 NCU admission born to adult pregnant women age 20-24 years old. Respiratory cases ranked first with 44(67.7%) cases, 11 (16.9%) with infectious in etiology, 2 (3.1%) cardiac cases, 4 surgical cases and 4 others.

**Table 4.21.**
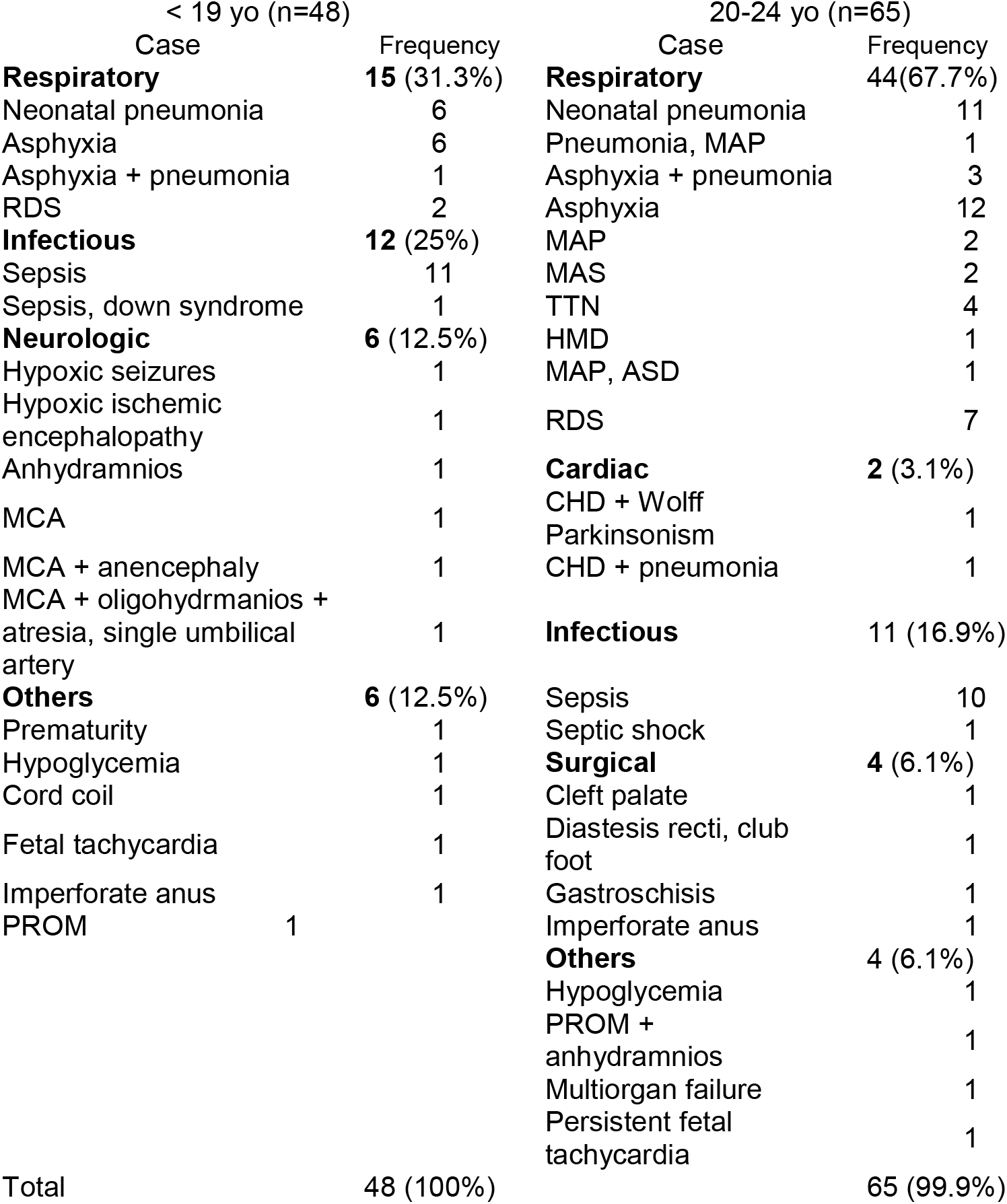
Prevalence of complications of NCU admitted babies born to teenage and 20-24 years old pregnant women at JRBGH from January to December 2017.

Tyrberg et al (2013), teenagers were more likely to deliver normally vaginally (aOR 1.70 (95%CI 1.64-1.75), less likely to have Caesarean section (aOR 0.61 (95%CI 0.58-0.64), and had a greater risk of delivering prematurely (< 28 weeks)(aOR 1.61 (95%CI 1.31-2.00), but did not have more small for-gestational-age babies (aOR 1.07 (95%CI 0.99-1.14). Furthermore, infants born to teenagers and women at advanced age possess greater risks for stillbirth, preterm birth, neonatal death, congenital anomaly, and low birth weight (Weng et al, 2014).

Objective No.3. To determine the cost profile (unit financial cost) of hospitalization of teenage pregnant women and the control group (based on Statement of Acount, SOA):

g Room and board fees
h Medicine/Supplies
i Laboratory
j Professional fee
k Delivery room/ Operating room fees
l Total hospital charges
m Philhealth benefit
n Patient’s hospital bill
o Medicare Type

The Statement of Account (SOA) presented the mean total hospital bill were Php 11,867.7 and 14,203, for teenage and 20-24 years old pregnant women, respectively. The total hospital bill is divided into the following, as follows: room fees, medicines used, laboratory fees, delivery room fees, Physcians’ professional fees. Table 4.22 above presents the mean expenses of pregnant women who were admitted in the facility. And in all items, (eg. room, medicines used, laboratory, delivery room and physicians’ professional fees), the mean fees were higher in the age group 20-24 years old compared to the teenage pregnant women.

**Table 4.22.** The cost profile (unit financial cost) of hospitalization of teenage pregnant women and 20-24 years pregnant women based on Statement of Accounts (SOA).

| Mean (Php) | < 19 yo (n=88) | 20-24 yo (n=91) |
| --- | --- | --- |
| Room Fee | 2,365 | 2,607 |
| Medicine | 3,989 | 5,461.8 |
| Laboratory Fee | 614.6 | 743.8 |
| Delivery Room Fee | 1,297.6 | 1,344.4 |
| Physicians' Professional Fee | 3,540.9 | 3,725.5 |
| **Total Mean Expenses** | **11,867.7** | **14,203** |
| No-Balance Billing | Majority (86/88) | Majority (73/91) |
| PhilHealth Benefits | 8,448.76 | 8,843.63 |

Based on the actual payment, majority of these patients availed the No-Balance Billing (NBB) policy of Philhealth and or the Medical Assistance Program (MAP) of the Department of Health as mandated by law. Private and self-employed individuals paid their total hospital bill but can also avail the MAP of DOH. Diagnosis which were considered as “case rates” were reimbursed by Philhealth, and the excess amounts will be funded by the MAP and not paid by the patients themselves.

All teenage pregnant women availed the NBB, except for 1 self-employed and 1 privately-employed (availed MAP). The average Philhealth benefits among these group was Php 8,448.76. Only 2 individuals paid the total hospital bills, and funded by Philhealth (Php=0).

Seventy-three (73) of 20-24 years old women availed the NBB. There were 18 women who were billed: 9 privately-employed and 9 self-employed (1 availed MAP). Thus, the average Philhealth benefits among these group was Php 8,843.63. Table 4.23, showed the distribution of those who were billed from ther hospital admission (bold letters).

**Table 4.23.** Distribution of teenage and 20-24 years pregnant women as to type of Medicare membership.

| Mean (Php) | < 19 yo<br>(n=88) | 20-24 yo<br>(n=90) |
| --- | --- | --- |
| LGU-Sponsored* | 34 | 29 |
| Indigent | 20 | 15 |
| POC** | 15 | 17 |
| POS*** | 13 | 10 |
| **Private Employed** | **1** | **9** |
| **Self-employed** | **1** | **9** |
| NHTS **** | 3 | 1 |
| 4P's ***** | 1 | 0 |
| Total | 88 | 90 |

Legend: \*LGU-Sponsored - local government unit-sponsored
\*\*POC - point-of-care (PhilHealth)
\*\*\*POS - point-of-service (PhilHealth)
\*\*\*\*NHTS - National Housing (DSWD)
\*\*\*\*\*4P's - Pantawid Pamilya (DSWD)

There were 88 (88/120, 73.3%) teenage pregnant women with SOA while 91 (91/131, 68.7%) from the 20-24 years old women. Based on the type of medicare, Table 4.23 demonstrated that most women in both groups were LGU-sponsored (local goverment unit-Cagayan De Oro City), 34 and 29 for teenage pregnant women and 20-24 years old women, respectively. Followed by 20 indigents, 15 for POC, 13 for POS, 3 were NHTS registered, and 1 each with membership from 4Ps, privately employed and self-employed, for the teenage group.

Moreover, the 20-24 year old women comprises with 17 POC, 15 for indigency program, 10 for POS, 9 both private and self-employed, and 1 for NHTS while none for the 4Ps membership.

## V. DISCUSSION

The issue of adolescent fertility is important for both health and social reasons. Children born to very young mothers are at increased risk of sickness and death. Teenage mothers are more likely to experience adverse pregnancy outcomes and to be constrained in their ability to pursue educational opportunities than young women who delay childbearing (PSA and ICF, 2018).

Moreover, 18% of teenagers in the Davao region and 15 percent each in Northern Mindanao and SOCCSKSARGEN have begun childbearing. Teenagers in the highest two wealth quintiles (3-5%) start childbearing later than those in other quintiles (10-15%) (PSA and ICF, 2018). Our local community contributed to this problem and in both perspectives, the mother (teenage) and the babies are affected in relation to health care.

Thus, searching for the knowledge and contributing for bridging the knowledge gap with special focus on adolescents’ sexuality and reproductive health, it has been percieved at the Unified Health Research Agenda (RUHRA) to redirect relevant solutions to the problems at the regional level through research. And teenage pregnancy is one of the reserach topics specified to achieved the objectives of NUHRA 2017-2022 (RUHRA CAR 2017-2022). The following statements assessed the hypotheses to statistical analysis for significant differences among variables (eg demographic profiles, clinical outcomes, and LOS) between the teenage pregnant women admitted at JRBGH and the 20-24 years old as the reference group for analysis.

### A. Maternal profile

Ho_1_: There is no significant difference between the <u>maternal profile</u> variables (residence, menarche, parity, blood pressure, timing of first ultrasonography, and mode of delivery) of teenage group vs control group pregnant women.

*HA*_*1*_: *There is a significant difference between the <u>maternal profile</u> variables (residence, menarche, gravidity, blood pressure, timing of first ultrasonography, and mode of delivery) of teenage group vs control group pregnant women*.

The mean age ang 120 teenage pregnant women is 17.29 years old (SD=1.3866) while 21.99 years old (SD=1.3352) for the 131 20-24 years old women. Among 8 demographic data from study pregnant women, those with significant diffrence with the two groups were the following; age, parity and timing of ultrasonography (Table 5.1).

**Table 5.1.** Test of difference (Z-test) between teenage vs young adults pregnant women (α =0.05).

| Pregnant Mothers Profile | $\leq 19$ yo (%) | 20-24 yo (%) | Z computed | P value |
| --- | --- | --- | --- | --- |
| Age (years), n | **120** | **131** |  |  |
| Mean | 17.29 | 21.99 | **-27.3493\*** | **0.00001** |
| +SD | 1.3866 | 1.3352 |  |  |
| Residence (n) | **119** | **130** |  |  |
| Urban | 77 (64.7) | 86 (66.2) | 0.2484ns | 0.8103 |
| Rural | 42 (35.3) | 44 (33.8) |  |  |
| Menarche (years) (n) | **96** | **102** |  |  |
| <13 yo | 69 (71.9) | 66 (64.7) | 1.0872ns | 0.283 |
| >14 yo | 27 (28.1) | 36 (35.3) |  |  |
| Parity (n) | **104** | **110** |  |  |
| 1 | 87 (83.7) | 65 (59.1) | **3.9638 \*** | **0.001\*** |
| >2 | 17 (16.3) | 45 (40.9) |  |  |
| Timing of ultrasonography(n) | **72** | **81** |  |  |
| 1st trimester | 0 (0) | 4 (4.9) | **2.7510\*** | **0.006\*** |
| 2nd trimester | 16 (22.2) | 31 (38.3) |  |  |
| 3rd trimester | 56 (77.8) | 46 (56.8) |  |  |
| Blood pressure (n) | **97** | **102** |  |  |
| Hypertensive | 11 (11.3) | 16 (15.7) | 0.9051ns | 0.3709ns |
| Non-hypertensive | 86 (88.7) | 86 (84.3) |  |  |
| Mode of delivery (n) | **119** | **129** |  |  |
| NSD | 95 (79.8) | 97 (75.2) | 0.8653ns | 0.3828ns |
| CS | 24 (20.2) | 32 (24.8) |  |  |
| Hepatitis B status (n) | **91** | **101** |  |  |
| Reactive | 1 (1.1) | 4 (4.0) | -- | -- |
| Non-reactive | 90 (98.9) | 97 (96.0) |  |  |

\* = significant difference

The incidence of teenage pregnancy ranges from 0.8% in Saudi Arabia (Abu-Heija et al, 2002) to as high as 27% (Pattanapisalsak, 2011); 4% Sagili et al (2012), 6% Kumar et al (2007), 5.10% Yasmin et al (2015), 7.1% Pambid (2015), 10% by Mahavarkar et al (2008) and Azevedo et al (2015), 13.3% Egbe et al (2015) and 15.24% from Thailand (Narukhutrpichai et al, 2016).

### 1. Residence

Kenyan study by Ochaku et al (2011) on young women aged 15-24 years old showed no significant difference based on the place of residence, either rural or urban.

The present study also showed no significant difference between the 2 groups either residing in an urban or rural residences. Both groups had residents from Barangay Carmen (most admission, urban) and the rural municipalities of Talakag, Baungon and Opol. Urban areas are strategically located and accessible by residents. Moreover, most of teenage pregnant women came from urban residences compared to the rural area. The same distribution in the 20-24 years old group.

In contrast, the current Philippine national survey showed that rural teenagers start childbearing slightly earlier than urban teenagers, 10% and 7% respectively (PSA and ICF, 2018). A hospital-based study in Egypt, showed that women were from rural areas (p=0.0001) among 606 teenage pregnant women versus 2,950 20-29 years old (Abbas et al, 2017).

### 2. Menarche

Among the 293 teenage mothers who gave birth in DLSUMC from January 2005-December 2009, 31.7% had their menarche at 12 years of age (Alon-Alon, 2010).

The present study presented the mean menarche of teenage pregnant women did not differ significantly compared to the 20-24 years old group. Moreover, the mode and median menarche (years) for each group is similar at 12 and 13 years old, respectively.

### 3. Parity

As expected in the present study, majority of the 104 teenage pregnant women were primigravida (83.7%) and lower in 20-24 years old (66, 64.7%). The result of the study showed a significant difference comparing the teenage and 20-24 years old pregnant women. There were 17 (16.3%) with repeat pregnancies among the teenage pregnant women. The maximun parity was 4 and 5 for the teenage and young pregnant women, respectively.

Comparable hospital-based studies were done by Khumar et al (2007) and Khier et al (2017). From the study of Khumar et al (2007), it showed 17% of the study group had their ultrasound done on their second or third trimester of pregnancy. The study consisted larger group of 369 pregnant women divided into 2 subgroups; ≤ 17 yr (group A) and 18-19 yr (group B). Meanwhile, Khier et al (2017) covered a total of 200 neonates (100 cases and 100 control group). This study showed significant differences between the two group (p value = 0.001) with most of the 19 years old group 63 (63%) and the majority of the >19 years old group 78 (78%) had number of parity between 1-4.

Contrary to the current and above studies, adults mothers were more parous than the teenage mothers (p=0.0001) (Abbas et al, 2017).

Succeding studies investigated the relationship of parity with sociodemographic variables and neonatal outcomes. Results of binary logistic regression by Singh et al (2014), parity is one of the signifiant determinants of the utilization of antenatal care services among urban married adolescent mothers (13-19 years). Highly significant advantages of low parity is shown with compliant and complete 4 ANC visits (*p <* 0.01), safe delivery (*p <* 0.01) and completed 2 post-natal care (*p <* 0.01).

A study investigated neonatal outcomes of all singleton teenage pregnancies with a study population consisted of 1,131 women; 75 12–15 years old, 163 16–17 years old, 157 18– 19 years old, and 736 24–29 years old. Parity is controlled and a significant linear association was found between maternal age and preterm birth, low birth weight, and neonatal birth related trauma (Suciu et al, 2014).

### 4. Timing of ultrasonography

Majority of the 20-24 years old group in the present study had their ultrasonography earlier (1st trimester, 4.9% and 2nd trimester, 38.3%) compared to the teenage group, 22.2%. This is shown to be statistically significant between the 2 groups. While most of the teenage group (77.8%) had thier UTS in the 3rd trimester, only half of the 20-24 years old group did their UTS at the last trimester.

Study by Khier et al (2017) from Sudan showed that 90 (90%) of the control group (>19 years old) and 88 (88%) of the case group (<19 years old) had ultrasound during pregnancy with no significant differences were between the two groups (P value = 0.162).

### 5. Blood pressure

The prevalence of hypertension (systole or diastole) in thepresent study is lower in teenage pregnant women (11.3%) than in 20-24 yeras old group (15.7%) and this is not statistically significant.

Complications like pregnancy induced hypertension (PIH) (11.4% vs 2.2%, p<0.01), pre-eclamptic toxemia (PET) (4.3% vs 0.6%, p<0.01) and eclampsia (4.9% vs 0.6%, p<0.01) occurred more commonly in teenagers compared to control (Kumar et al 2007). But not to the group studied by Kovavisarach et al 2010 among those with mild pre-eclampsia (p-value 0.774).

### 6. Mode of delivery

Regarding the mode of delivery in the two groups, normal delivery was 48 (48%) in control group and 46 (46%) in cases; instrumental 7 (7%) in control and 2 (2%) in cases; 45 (45%) of the control group and 52 (52%) of the cases delivered through caesarean section with no significant difference between the two groups (P value = 0.069) according to the study by Khier et al (2017) and Gilany and Hammad (2012). The incidence of caesarean section in low teens among Iranians (<16 years old) was higher than high teens (17-19 years old (P=0.001), but caesarean was not higher among mothers aged 19 years or less compared with women aged 20-29 years, according to Khooshideh and Ali (2008).

Tyrberg et al (2013) noted that teenagers were more likely to deliver normally vaginally (aOR 1.70 (95%CI 1.64-1.75), less likely to have Caesarean section (aOR 0.61 (95%CI 0.58-0.64). In contrast, cesarean delivery in the teenage group was significantly higher than that in the adult group (18.7%: 13.3%, p = 0.006), according to Kovavisarach et al 2010. In addition, vacuum extraction in the teenage group was significantly lower than that in the adult group (0.8%: 2.8%, p = 0.006).

### 7. Hepatitis B

The present study showed only one (1.1%) tested Hepatitis B reactive mother and majority were non-Hepatitis B reactive (90, 98.9%) among the teenage group. This was not subjected to statistical analysis due to small subject population. Moreover, 20-24 years old group, same trend was noted, majority were non-Hepatitis B reactive (97, 96%) and there were 4 (4%) tested Hepatitis-B reactive.

The risks for hepatitis B and hepatitis C, were seen lower in teenage pregnant women compared with those in the young adult group, respectively, 0.43 (CI 0.26–0.71), 0.90 (CI 0.85– 0.96), according to Socolov et al (2017). In addition, subgroups within teenage pregnant women showed consistent and significantly high Hepatitis B infection (in Romania): age 12–17 years old = 3 (0.23%); 18-19 years old = 15 (0.57%); 12–19 =18 (0.46%); at p-value< 0.01.

### 2. Length of hospital stay (mother)

Ho_2:_ There is no significant difference between the mean <u>length of hospital stay</u> among teenage group versus the control group of pregnant women (20-24 yo).

*H*_*A2:*_ *There is a significant difference between the mean <u>length of hospital stay</u> among teenage group versus the control group of pregnant women (20-24 yo)*.

This study concluded that there is no significant difference between the mean length of hospital stay (LOS) among teenage group(5.16) versus the control group (5.54) of pregnant women (20-24 yo). Fewer studies had similar conclusion with the present study by Socolov et al (2017) and Suciu et al (2015).

According to Socolov et al (2017), the length of hospital stays for parturients was not significantly different between the 2 age groups: 6.44 ± 3.89 days for teenagers versus 6.59 ± 4.23 days for young adults (= 0.06) and by Shah et al (2011) 1.6±2.2 days and 1.9±2.3days (mean±SD) for teenage women and adult women, respectively. Moreover, using 395 teenage pregnant women segregated into early (13–16 years), late teenage (17–19 years), and adult (25–29 years), the length of hospital stay was similar with adult mothers’ newborns (Suciu et al, 2015).

There are several published studies showing that LOS is longer in teenage pregnant women. Adebanji et al (2015) studied 180 singleton mothers at Sunyani regional and municipal hospitals. It concluded that teenage mothers were associated with longer LOS (mean=7.61 ± 1.147) compared to middle age mothers (mean=6.27 ± 1.225).

Results of the study by Suciu et al (2014) stated that length of hospitalisation, as a marker of the healthcare costs involved in the care of these high risk cases, was significant associated with maternal age. Furthermore, it concluded that teenage pregnancy is a risk factor for poor neonatal outcome (low birth weight, preterm delivery, neonatal birth related trauma) and high healthcare costs among 1,131 pregnant women.

Ironically, the mean length of stay for admissions related to pregnancy among adolescents was 2.6 days for both studies by Fingar and Hambrick (2013) and Areemit et al (2012). Greater percentage of childbirth stays paid by Medicaid was nearly 2 times higher among teens (majority, 72.3 % by 18-19 years old) than among women aged 20–44 years (Fingar and Hambrick, 2013) and the average cost per stay was $4,300. Results of Areemit et al (2012) was in contrast to the above study, which was significantly less than that of women in the optimum reproductive age (2.8 days) compared to the 2.6 days.

### 3. Mean hospital expenses (mother)

Ho_3_: There is no significant difference in the <u>mean hospital expenses</u> between teenage vs the 20-24 years old pregnant women whose babies were admitted at NCU.

*H*_*A3*_: *There is a significant difference in the <u>mean hospital expenses</u> between teenage vs the 20-24 years old pregnant women whose babies were admitted at NCU*.

The mean hospital expenses for the teenage pregnant women was lower at Php 11,868 while Php 14,203 for the young adult pregnant women, p value=0.01986. Test of mean difference showed a significant difference in the mean hospital expenses between the two groups. Using the converison of USD1 to Php51, the mean hospital expenses would be $232.7 and $278.5 for teenage and young adult pregnant women, respectively (Table 5.3).

**Table 5.2.** Test of difference of two means of length of hospital stay between teenage vs young adults pregnant women (α =0.05).

| Parameter | ≤19 yo | 20-24 yo | p-value | Mean difference | Lower limit | Upper limit |
| --- | --- | --- | --- | --- | --- | --- |
| Length of stay (days) | 98 | 107 |  |  |  |  |
| Mean | 5.16 | 5.54 | 0.3411<sup>ns</sup> | -0.38 | -1.16525 | 0.40525 |
| Standard deviation | 2.455348 | 3.165454 |  |  |  |  |

ns = no significant difference, t-test t = -0.954165 > Critical region t = -1.96, not within the critical region

**Table 5.3.** Test of difference between two means of hospital expenses between teenage vs young adults pregnant women (α =0.05).

| Parameter | ≤19 yo | 20-24 yo | p-value | Mean difference | Lower limit | Upper limit |
| --- | --- | --- | --- | --- | --- | --- |
| Hospital expenses (n) | 88 | 91 |  |  |  |  |
| Mean | 11,867.7 | 14,203 | 0.01986* | -2335.3 | -4296.11 | -374.492 |
| Standard deviation | 5,175.7 | 7,808 |  |  |  |  |

\* = significant difference
t= -2.35036 < Critical region t=-1.96, within the critical region

Correlating the LOS and expenses, Fingar and Hambrick, (2013) reported that the mean length of stay (2.6 days) would be equivalent to $4,300 as an average cost per stay. Greater percentage of childbirth stays paid by Medicaid (in the US) was nearly 2 times higher among teens (majority, 72.3 % by 18-19 years old) than among women aged 20–44 years. In 2013, the cost of teen hospital stays for childbirth amounted to over $1.1 billion.

From the 84 neonates conducted by Mwamakamba and Zucchi (2014), they had a total of 1,633 days in-patient hospital care at a total cost of US$195,609.00. Approximately 72% of this total cost (US$141,323.00) accounted for hospital services. The mean daily costs ranged from US$97.00 to US$157.00

Another hospital-based study in city of Sao Paulo, correlated the cost to the respective weight of the neonates, namely: <1000g, 1000-1,499, 1,500-1,999, and >2,000 g. For neonates weighing <1,000g and from 1,000 to 1,499g, the mean hospital stay of 34 and 42 days, with average neonatal costs of US$3,919.00 and US$4,043.00, respectively. For those weighing from 1,500 to 1,999g and >2,000 g, the average cost ranged from US$1,768.00 to US$1,183.00, and the total direct cost of treatment was US$2,017.00 (Desgualdo, 2011).

The mean hospital charges per admission and cumulative cost for admissions related to pregnancy among adolescents was 5,723 Baht and 830.4 million Baht, which was less than for women in the optimum reproductive age group (*viz*., 6,744 Baht and 2,635.4 million Baht). Though less, it was without statistical significance in the 10-14 age group and with statistical significance in the15-19 age group, because data in the10-14 age group had more deviation (Areemit et al, 2012).

This hospital-based study in JRBGH, does compute for the total days in-patient relative to the total number of NCU in ptients because some SOA cannot be retrieved.

### 4. NCU profile variables (Neonates)

Ho_4_: There is no significant difference and association between <u>NCU profile variables</u> (gender, birth weight, Apgar score, and outcome) born to teenage group vs control group pregnant women.

*HA*_*4*_: *There is a significant difference between NCU profile variables (gender, birth weight, Apgar score, and outcome) born to teenage group vs control group pregnant women*.

Present study showed no significant difference between the two groups in terms of sex, birthweight, Apgar score and clinical outcome. Several studies also presented no significant diffrence between teenage pregnancy and young adults as reference groups (Tyrberg et al 2013, El-Gilany and Hammad, 2012).

Several studies concluded that teenage pregnancy is associated with poor neonatal outcomes, namely: increased morbidity and mortality rates, prematurity, preterm deliveries, low birth weight (LBW), and poor Apgar score. This study is focused on the last two clinical problems, poor Apgar score (1st 1 minute) and LBW as variables relative to two age group, namely; teenage pregnancy and young adult pregnant women. (20-24 years old).

Chen et al (2007), showed that the rates of very LBW, LBW, very low Apgar score, low Apgar score and neonatal mortality were higher in teenage pregnancies. Moreover, these rates and risks were consistently increased with decreasing maternal age and were always highest among infants born to mothers aged 15 years or younger than infants born to mothers of 20–24 years old.

A large study population covered 4,254,751 first-born singleton infants: 0.85% infants born to younger teenage mothers aged 10–15 years old, 3.02% to women aged 16–17 years and 4.89% to women with 18–19 years old were used in the study by Chen et al (2007).

Teenage mothers subdivided into 2 groups, showed that ≤17 years were most vulnerable to poor neonatal outcome compared to 18-19 years mothers (Kumar, 2007). They also had increased incidence of low birth weight (LBW) (50.4% vs 32.3%, p<0.01) and were associated with higher fetal (1.9% vs 0.3%, p<0.05) and neonatal mortality (3.8% vs 0.5%, p<0.05).

Nili et al (2002) evaluated 102 neonates from 99 women under 18 years during April 1999 to April 2000. Study showed that 32% of neonates were low birth weight (LBW) and the gestational age in 38.2% of neonates was lower than 37 weeks. Neonatal mortality occurred in 6.9% of patients.

Published studies either to compare the teenage with the young adults pregnant women or subgroup the teenage population(18-19 yo, 16-17 yo etc), majority of these studies documented the increased risks and rates of low birth weights: Chen et al (2017); Khier et al (2017) p-value 0.001; Kovavisarach et al (2010) p-value <0.001; 87.20% (Khumar, 2007); OR=1.8 p<0.0001 (Mahavarkar et al, 2008); OR 1.9 (Pattanna, 2011); Ara and Alam (2017) p-value <0.05 and Thobbi et al (2017) 20%; review of related literatures also cited studies from Chirayu and Chandeying (2012); Abu-Heija et al (2002), p<0.001, and significantly high LBW among teenage pregnant women (Sagili et al,2012).

But some studies concluded low rates of LBW from teenage pregnant women: Lama et al (2015) p-value 0.259; Pun and Chauhan (2011); Suwal (2012) and Egbe et al (2015).

This present study and Tyrberg et al (2013) showed that Apgar score was similar to the reference group, while some studies showed significant difference between the 2 groups of pregnant women Ara and Alam (2017), Derme et al (2013), and Kovavisarach et al, (2010).

Tyrberg et al (2013) subdivided adolescents into three groups: <16 years (n = 472), 16– 17 years (n = 5376), 18–19 years (n = 23,560). This study showed the rate Apgar score <7 at 5 minutes was similar to the reference group consisted of women age 20–30 years (n = 893,505). This low Apgar score (p-value 0.377) is duplicated by Khier et al (2017).

Apgar score showed significant difference between the two groups in study by (Ara and Alam (2017). Twelve percent neonates of teenage mothers were severely asphyxiated (Apgar score 0 to 3), which was significantly higher than control group (4%). At 1 minute, 52.0% babies of teen mothers have more than 7 Apgar score, whereas 86.0% babies born to non-teen mothers showed an Apgar score more than 7 which was also a significant difference

Majority of the newborns in the study group presented a normal APGAR score at the first and fifth minute after birth (index of wellbeing of the baby): the APGAR score at the first minute ≥7 for 170 babies (94.4%); the Apgar score at the fifth minute ≥7 for 178 babies (98.9%). In the control group we observed the Apgar score at the fifth minute ≥7 for 139 babies (92.7%) (p=0.004) (Derme et al, 2013).

Kovavisarach et al, 2010 presented significant difference of Apgar score (first 1 min, <7) between 750 teenage mothers compared to 750 adult mothers (p-value= 0.014, OR 2.05, 1.17-3.60). There was no significant difference between the two age groups at Apgar score (<7 at 5 minutes), p-value= 0.050.

On the other hand, neonatal outcomes did not differ between teenage mothers (<20 years) and the reference group (20-34 years) using a retrospective record-based comparative study in Saudi Arabia (El-Gilany and Hammad, 2012). This present study subdivided the neonatal outcomes in terms of complicated and non-complicated which were not use in the study of El-Gilany and Hammad (2012).

### 5. Length of NCU stay (Neonates)

Ho_5_There is no significant difference between the mean <u>length of NCU stay</u> among babies born to teenage group versus the control group of pregnant women (20-24 yo).

*Ho*_*5:*_ *There is a significant difference between the <u>mean length of NCU stay</u> among babies born to teenage group versus the control group of pregnant women (20-24 yo)*.

Table 5.5, in this study, showed no significant difference between the mean length of stay (LOS) at the NCU of newborn delivered by the teenage (3.38 days) and the 20-24 years (3.23 days) old pregnant women (p-value=0.8138).

**Table 5.4.** Test of difference (Z-test) of newborn profile born to teenage vs young adults pregnant women (α =0.05).

| Newborn Profile | ≤19 yo (%) | 20-24 yo (%) | Z computed | P value |
| --- | --- | --- | --- | --- |
| Sex (n) | 120 | 131 |  |  |
| Male | 74 (61.7) | 83 (63.4) | -0.00001ns | 0.7820 |
| Female | 46 (38.3) | 48 (36.6) |  |  |
| Birth weight (n) | 124 | 131 |  |  |
| ≤ 2.4 kgs | 53 (42.7) | 41 (31.2) | 1.9026ns | 0.0583 |
| ≥ 2.5 kgs | 71 (57.3) | 90 (68.8) |  |  |
| Apgar score (n) | 121 | 131 |  |  |
| ≤ 6 | 42 (34.7) | 49 (37.4) | -0.4458ns | 0.6565 |
| ≥ 7 | 79 (65.3) | 82 (62.6) |  |  |
| Outcome (n) | 109 | 120 |  |  |
| Complicated | 23 (21.2) | 23 (19.2) | 0.3772ns | 0.7152 |
| Non-complicated | 86 (78.8) | 97 (80.8) |  |  |

\* = significant difference, z test

**Table 5.5.** Test of difference between two means of newborn NCU stay (days) born to teenage vs young adults pregnant women (alpha= 0.05).

| Parameter (NCU days) | ≤19 yo | 20-24 yo | p-value | Mean difference | Lower limit | Upper limit |
| --- | --- | --- | --- | --- | --- | --- |
| Length of stay (n) | 105 | 120 |  |  |  |  |
| Mean | 3.38 | 3.23 | 0.8138<sup>ns</sup> | 0.15 | -1.10373 | 1.40373 |
| Std. Dev. | 5.6678 | 3.7948 |  |  |  |  |

ns = not significant difference P value> alpha 0.05 t= 0.213 < Critical region t=1.96

Findings of the current study is similar with Adebanji (2015), irrespective of birth-status and gestational age. Meanwhile, published study by Mishra et al (2005) associated longer LOS to LBW neonates, but the study population included mothers ages > 20 years old.

Newborn LOS reflects the underlying health condition at birth, and the risk of readmission tended to increase with longer newborn LOS, according to Harron et al (2017). Furthermore, the median newborn length of stay was 1 day (2 days for late preterm babies). Overall, 5.2% (n = 244,827) of babies in the study population had one or more unplanned readmissions within 30 days post-discharge (7.2% for early term, 10.6% for late preterm births). The study population was not compared specifically with the teenage mothers.

The length of stay of 84 neonates (specifically, preterm) had a median length of stay of 19.44 days; and varied average days in terms of birthweight: <1,000g, 32.25 days; 1,000-1,499g, 26.0 days;1,500-1,999g, 5.93 days; and >2,000g, 1.57 days (Mwamakamba and Zucchi, 2014).

## VI. CONCLUSION

Though most studies had increased rates and risks of maternal and neonatal morbidities and mortalities, this study showed no ultrasound performed for the first trimester on high risks teenage pregnant women. The first trimester ultrasonography is considered vital in health and care of both the mother and the baby. Possible and varied reasons may existed (eg financial, delayed confirming of the unwanted pregnancy) but beyond the scope of this study. However, it is shown in the study that more teenage pregnant women had their UTS in the 3rd trimester (late) and more 20-24 years old had their UTS in the first and second trimester (earlier) which is statistically significant.

The projected population in Cagayan de Oro City is increasing and this is reflected in an increassing yearly census of deliveries in JRBGH (Figure 4.1). These may not be felt as burden to the family because of PhilHealth financing, but the LGU, as a stakeholder, and as health financing agency, may be affected in delivering services to these age groups and to other LGU departments which needs more prioritization of funds.

## VII. RECOMMENDATION

The proponent of the study recommends the following:

1. To improve data collection for the hospital-based collection and community surveys to maintain a large sample size in each variables to be use.
2. More variables should be utilized to widened the scope of assessing these teenage pregnant women.
3. Teenage pregnant women may be covered in other areas of the communities like the school entrants, out-of-school youth and hospital admissions (public and private hospitals) and health centers.
4. Improved ante-natal care by focused-group discussions among teenage pregnant women only and emphasize post-natal care visits.
5. Collaboration of local stakeholders with the teenage programs, namely: POPCOM, DepED, City Health Office, JRBGH, LGU, PPS, and DSWD.
6. Good tracking system or notebooks carried by each teenage pregnant women for better referral system and data collection (for analysis).
7. To conduct similar study with a larger sample size and find more association among maternal and neonatal profiles.

## Data Availability

All data produced in the present work are contained in the manuscript

